# Use of Personal Protective Equipment in General Practice and Ambulance settings: a rapid review

**DOI:** 10.1101/2023.03.10.23287113

**Authors:** Antonia Needham, Tom Winfield, Lauren Elston, Jenni Washington, Ruth Lewis, Alison Cooper, Adrian Edwards

## Abstract

The use of personal protective equipment (PPE) is a cornerstone of infection prevention and control guidelines and was of increased importance during the COVID-19 pandemic. Adherence with prescribed guidelines for the use of PPE and their applicability to the working practices of staff in general practitioner (GP) and ambulance settings have been a growing concern. The aim of this rapid review was to assess the barriers, facilitators, and potential adverse outcomes of the use of PPE in these specific settings.

Included studies were published from 2020 to November 2022. We identified four systematic reviews, a rapid review, a retrospective chart review, and a prospective quantitative survey study. Outcome measures were broadly focused on physical adverse outcomes from the use of PPE, but also included barriers and facilitators to the use of PPE in varied healthcare settings. The five reviews covered a broad range of health and care settings, which included GP and ambulance settings, but not as a specific focus. Both the retrospective chart review and the prospective survey study took place in an ambulance or emergency response setting. Overall confidence in the body of evidence is low.

Extended use of PPE is associated with an increased occurrence of adverse physiological events, such as pressure ulcers and de novo headaches. Evidence indicates that adherence with PPE guidance is primarily influenced by organisational communication and workplace cultures. In ambulance settings, adherence may also be affected by dispatch codes and indicative symptoms reported during the initial call.

Policy implications: As there is evidence to suggest that usage of PPE increases risk of adverse effects in healthcare workers, this should be at the forefront of considerations when developing or reviewing new and existing infection prevention and control measures. If new policy regarding the use and implementation of PPE is to be developed, effective communication and dissemination should be a priority, as this was identified as a barrier to adherence. This review has identified a significant paucity of evidence in the settings of interest and is reliant on examining evidence that represents a large variety of health and care settings. It is important to acknowledge there may be some issues specific to Ambulance and GP settings that are not covered by this review. This does impact the validity of this reviews conclusions.

Further high-quality research must be undertaken in the settings of interest to inform and guide policy.

**Funding statement:** Health Technology Wales was funded for this work by the Wales Covid-19 Evidence Centre, itself funded by Health & Care Research Wales on behalf of Welsh Government.

**Rapid Review Details:** *Review conducted by:* Health Technology Wales (HTW)

*Review Team:*

- Antonia Needham, Health Technology Wales,
- Tom Winfield, Health Technology Wales,
- Lauren Elston, Health Technology Wales,
- Jenni Washington, Health Technology Wales

*Review submitted to the WCEC on:* 10th February 2023

*Stakeholder consultation meeting:* 23^rd^ January 2023 [day, month, year]

*Rapid Review report issued by the WCEC:* February 2023

*WCEC Team:* Adrian Edwards, Ruth Lewis, Alison Cooper and Micaela Gal were involved in drafting the Topline summary, review of the report and editing

*This review should be cited as:* RR00046. Wales COVID-19 Evidence Centre. Use of Personal Protective Equipment (PPE) in General Practice and Ambulance settings: a rapid review. February 2023.

*Disclaimer:* The views expressed in this publication are those of the authors, not necessarily Health and Care Research Wales. The WCEC and authors of this work declare that they have no conflict of interest.

**TOPLINE SUMMARY:** *What is a Rapid Review?:* Our rapid reviews (RR) use a variation of the systematic review approach, abbreviating or omitting some components to generate the evidence to inform stakeholders promptly whilst maintaining attention to bias. They follow the methodological recommendations and minimum standards for conducting and reporting rapid reviews, including a structured protocol, systematic search, screening, data extraction, critical appraisal, and evidence synthesis to answer a specific question and identify key research gaps. They take 1-2 months, depending on the breadth and complexity of the research topic/ question(s), extent of the evidence base, and type of analysis required for synthesis.

*Who is this summary for?:* Wales Ambulance Service NHS Trust and the Royal College of General Practitioners Wales

*Background / Aim of Rapid Review:* The use of personal protective equipment (PPE) is a cornerstone of infection prevention and control guidelines and was of increased importance during the COVID-19 pandemic. Adherence with prescribed guidelines for the use of PPE and their applicability to the working practices of staff in general practitioner (GP) and ambulance settings have been a growing concern. This rapid review aims to assess the barriers, facilitators, and potential adverse outcomes of the use of PPE in these specific settings.

*Key Findings:* Extent of the evidence base

- We identified four systematic reviews (Galanis et al, 2021; Keng et al, 2021; Kunstler et al, 2022), one rapid review (Houghton et al, 2020), a retrospective chart review (McCann-Pineo et al 2022) and a prospective quantitative survey study (Gangaram et al 2022).
- Outcome measures were broadly focused on physical adverse outcomes from the use of PPE, but also included barriers and facilitators to the use of PPE in varied healthcare settings.
- In terms of setting, all five systematic and rapid reviews covered a broad range of health and care settings, all of which included GP and ambulance settings, but not as a specific focus – it was deemed that as these settings were included as part of data collection and analysis that the findings would be generalisable.
- Both the retrospective chart review (McCann-Pineo et al 2022) and the prospective survey study (Gangaram et al 2022) took place in an ambulance or emergency response setting. Recency of the evidence base

- Studies included were published from 2020 up until November 2022. Key Findings

- There is a significant lack of evidence in the settings of interest.
- Extended use of PPE is associated with an increased occurrence of adverse physiological events, such as pressure ulcers and de novo headaches.
- Evidence indicates that adherence with PPE guidance is primarily influenced by organisational communication and workplace cultures. In ambulance settings, adherence may also be affected by dispatch codes and indicative symptoms reported during the initial call. Quality of the evidence

- Of the systematic reviews identified (Galanis et al, 2021; Keng at al, 2021; Kunstler et al, 2022) all are of poor quality, and were determined to have high risk of bias following formal assessment.
- The rapid review identified (Houghton et al, 2020) is of good quality, with a low risk of bias.
- Of the primary studies (McCann-Pineo et al, 2022; Gangaram et al, 2022) the retrospective chart review was deemed ‘poor’ quality with high risk of bias, and the prospective quantitative survey study deemed ‘fair’ quality, with undetermined risk of bias.
- Primary concerns around the evidence base relate to evidence identification, applicability of evidence and methodological limitations. Policy Implications

- There is evidence to suggest that usage of PPE increases risk of adverse effects in healthcare workers, and this should be at the forefront of considerations when developing or reviewing new and existing infection prevention and control measures.
- If new policy regarding the use and implementation of PPE is to be developed, effective communication and dissemination should be a priority, as this was identified as a barrier to adherence.
- This review has identified a significant paucity of evidence in the settings of interest and is reliant on examining evidence that represents a large variety of health and care settings. It is important to acknowledge there may be some issues specific to Ambulance and GP settings that are not covered by this review. This does impact the validity of this review’s conclusions.
- Further high-quality research must be undertaken in the settings of interest to inform and guide policy. Strength of Evidence
Overall confidence in the body of evidence is low, and caution should be exercised when drawing conclusions based on this evidence.

## 1. BACKGROUND

### 1.1 Who is this review for?

This rapid review was conducted as part of the Wales COVID-19 Evidence Centre (WCEC) Work Programme. The above question was suggested by both Wales Ambulance Service NHS Trust and the Royal College of General Practitioners. The research question was developed through collaboration with a range of stakeholders, including the WCEC Core Team, Health Technology Wales, Welsh Ambulance Services NHS Trust, Royal College of General Practitioners Wales, and public contributors.

### 1.2 Background and purpose of this review

During the COVID-19 pandemic, the transmission of SARS-CoV-2 in healthcare settings has been of great concern. This has resulted in the development of enhanced infection prevention and control (IPAC) guidelines, of which the use of personal protective equipment (PPE) forms a significant part. How these PPE guidelines were to be used, and how individual members of staff across various settings and roles adhere to them were of growing concern over the course of the pandemic due to a need to improve safety for healthcare workers and allied staff, as well as the patients in their care. Early IPAC guidelines and regulations were developed primarily with the hospital setting in mind, leading to uncertainty about their provision and use in other health and care settings. The purpose of this review is to evaluate the barriers, facilitators and adverse outcomes of the use of PPE in specific GP and ambulance settings.

Primary health care workers, such as those in the GP and ambulance environment, are at the forefront of infection management for all respiratory diseases, a responsibility that has increased dramatically during the COVID-19 pandemic. Staff in these settings are at an increased risk of infection and transmission due to their increased exposure to symptomatic and asymptomatic people and the number of aerosol-generation procedures that occur in the ambulance environment. In these settings, the level of coverage and specific type of PPE required is primarily determined by level of risk, measured by the number of Aerosol-Generating Procedures (AGPs) that occur. However, the primary types of PPE recommended in these settings are face coverings, gloves and gowns, and it is these categories that this review will consider.

## 2. RESULTS

### 2.1 Overview of the Evidence Base

We searched for published and pre-print articles where barriers, facilitators and adverse outcomes of PPE were discussed. As per the research protocol, systematic and rapid reviews were prioritised in order to conduct a ‘review of reviews’. The search was not restricted to COVID-19, including other related respiratory viruses such as MERS-CoV and influenza and publication dates were limited between 2020-2022. We identified four systematic reviews and one rapid review. All five reviews reported on healthcare settings more generally, including the settings of interest for this review (GP and ambulance settings), but did not report evidence on these settings separately. We therefore searched and included any primary evidence that reported on GP or ambulance settings specifically. Two primary studies were identified, both in the ambulance or emergency response setting. We did not identify any evidence that reported on GP settings specifically.

### 2.2 Summary of the evidence

#### 2.2.1 Systematic Reviews

We identified four systematic reviews in this rapid review (Galanis et al 2021, Keng et al 2021, Kunstler et al 2022, and Tezcan et al 2022).

Galanis et al (2021) conducted a systematic review and meta-analysis on the impact of PPE in health care workers’ physical health during the COVID-19 pandemic with an aim to identify factors that related to a greater risk of adverse physical health outcomes stemming from PPE use. A total of 14 studies were included in the review, with a total sample size of 11,746 health care workers from a variety of settings. Of the studies included, 11 investigated risk factors for adverse events, six used multivariable models to eliminate confounding factors and all studies bar one measured the occurrence of any adverse event as the dependent variable. The review found that prevalence of any adverse event ranged from 42.8% to 95.1% among individual studies and the most common adverse events reported were dry skin, pressure injuries, headache, and dermatitis. The overall pooled prevalence of adverse events was 78% (95% confidence interval [CI] 66.7% to 87.5%). The review also explored risk factors for adverse events that were reported by individual studies; certain demographic, clinical and job characteristics were related to increased risk of adverse outcome such as duration of shift wearing PPE, and use of higher grade PPE.

Keng, et al (2021) conducted a systematic review of occupational dermatoses related to PPE and their prevalence among health care workers. The objective of the review was to determine the most common types of PPE-related dermatoses, the affected body sites, and occupational risk factors for their development. Sixteen studies were included in this review, with a total sample size of 3,958 health care workers in diverse environments. The review found that the most prevalent adverse outcomes identified in the overall sample size (as opposed to most commonly reported within each study) were xerosis (27.6%), pressure-related erythema (22.1%) and irritant contact dermatitis (14.8%), with face and hands being the most commonly affected areas. The increased use of gloves and N95 masks were two of the most prevalent PPE measures implicated in the formation of these dermatoses (34.2% and 26.9%, respectively).

Kunstler, et al (2022) conducted a systematic review and meta-analysis on the effectiveness and adverse effects of P2/N95 respirators and surgical masks in the context of the COVID-19 pandemic. Twenty-one studies were included, with a total sample size of 11,744. The studies included were primarily observational studies, with one randomised controlled trial. The review found that in terms of adverse events, healthcare workers are significantly more likely to experience headache (odds ratio [OR] 2.62, 95% CI 1.18 to 5.81, 3 studies), respiratory distress or shortness of breath (OR 4.21, 95% CI 1.46 to 12.13, 3 studies), facial irritation (OR 1.80, 95% CI 1.03 to 3.14, 3 studies) and pressure related injuries (OR 4.39, 95% CI 2.37 to 8.15, 3 studies) when wearing respirators compared to surgical masks.

Tezcan, et al (2022) conducted a systematic review of PPE-related pressure ulcers in healthcare workers. Seventeen studies were included, for a total sample size of 24,889. The review identified that the rate of PPE-related pressure ulcers across the studies ranged between 30% and 92.8%. The most common sites for pressure ulcers were on the nose, ears, forehead, and cheeks. Similarly, while pressure ulcers reported varied in severity, grade one pressure ulcers, where the skin remains intact but non-blanchable redness occurs, was the most commonly reported type among those sampled.

#### 2.2.2 Rapid Review

One rapid review was identified. Houghton et al (2020) performed a rapid qualitative evidence synthesis of the barriers and facilitators to healthcare workers’ adherence to IPAC guidelines for respiratory infectious diseases. The review included 20 qualitative and mixed-methods studies that focused on healthcare workers’ experiences and views of IPAC guidelines for SARS, H1N1, MERS, tuberculosis, and seasonal influenza. The study identified several barriers and facilitators that influence adherence to guidelines, primarily focused on clear communication about guidelines and clarity when local and national/international guidelines are conflicting. Similarly, availability and provision of PPE was identified as a significant barrier. In terms of facilitators, the review identifies the importance of workplace culture and notes that healthcare workers believed they followed guidance more closely when they were able to see its value in providing quality care to patients.

#### 2.2.3 Primary Studies

Two primary studies were identified in this rapid review. One prospective quantitative survey study (Gangaram et al, 2022) and one retrospective chart review (McCann-Pineo et al, 2022) are included due to their specific focus on the ambulance or emergency medical services setting.

Gangaram et al (2022) performed a prospective quantitative survey study, collecting descriptive data from frontline paramedics employed by the Hamad Medical Corporation Ambulance Service in Qatar. One thousand paramedics were invited via email to complete an online survey, with a total response rate of 28.2% (282 paramedics). The study found that paramedics’ attitudes toward PPE are primarily positive but identified some confusion over the process of donning and doffing PPE and how it integrates with standard practice. The study also noted the HMCAS standard operating procedure of supervised doffing of PPE, with 80.1% of respondents reporting they practice this.

McCann-Pineo et al (2022) performed a retrospective chart review of factors influencing the use of PPE among EMS responders during the COVID-19 pandemic in New York City. All adult emergency encounters with available prehospital data that occurred between March 16 and June 30^th^, 2020, were analysed via multinomial logistic regression. The study identified 28,693 eligible encounters and found that PPE was used in 92.8% of them, with ‘full’ PPE in 17.8%. Full PPE utilisation, defined by the study as donning gloves, eye protection, face mask and a gown, was largely dependent on the dispatch code communicated to EMS. The study identifies that codes that were indicative of breathing problems or respiratory arrest influenced decisions around full PPE use.

#### 2.2.3 Bottom line summary

The evidence identified in this review suggests there are multiple adverse outcomes of PPE use, which are primarily physiological in nature. Outcomes such as pressure ulcers, de novo headache and dermatitis were commonly reported across multiple studies. One review identified the importance of workplace culture and values in facilitating the use of PPE, and how communication and clarity regarding perceived conflicts between local and international guidelines can affect adherence, becoming a barrier.

Across all studies identified in this review there is a clear paucity of evidence that is specific to the settings of interest. While generalisable themes have emerged from the reviews included, and the settings of interest have been represented in part, further high quality evidence is required to inform any decision making in this area.

## 3. DISCUSSION

### 3.1 Summary of the findings

In order to appropriately address the desired research questions the discussion of the evidence is split into three sections, covering adverse outcomes of PPE, barriers to PPE adherence, and facilitators to PPE adherence.

#### 3.1.1 Adverse outcomes of PPE

The majority of adverse outcomes stemming from the use of PPE that have been identified in this rapid review are dermatological in nature. Reviews such as Galanis et al (2021) identify that skin reactions are the most frequent adverse outcome of PPE as reported by HCWs – the study identifies that while gloves, filtering face piece respirators and hand sanitization are ‘indispensable’ in the control of COVID-19, their routine use may lead to a removal of normal bacterial flora and disruption of the skin’s protective barrier, potentially worsening the frequency and severity of skin diseases. Keng et al (2021) echoes this, finding that pressure-related skin injuries are a frequent complication of N95 masks and goggles, especially when worn for long period, and the presence of skin maceration and abrasions caused by PPE have the potential to progress to fissures, erosions, blisters or ulcers – if these abrasions are situated on the nasal bridge and cheeks, two of the most susceptible locations, there is the potential for a compromised skin barrier and a secondary infection may manifest. Similarly, the evidence suggests the potential for face coverings to cause or worsen skin conditions in HCWs; for example, the excessive accumulation of sweat and sebum that occurs due to increased heat and humidity, combined with increased friction and pressure from donning and doffing PPE may result in the manifestation of acne (Keng et al, 2021; Tezcan et al, 2022). The evidence also suggests that the occurrence of dermatological adverse effects may be greater in HCWs who wear respirators as opposed to surgical masks, though the evidence for this is limited (Kunstler et al, 2022).

Another adverse outcome identified by the literature is the occurrence of de novo headache. While pre-existing headache conditions place HCWs at increased risk for experiencing this outcome (Galanis et al, 2021) the evidence suggests that headaches are common among HCWs who wear filtering face piece respirators. Occurrence of headaches in these cases are positively associated with the length of time the face piece is worn (Galanis et al, 2021; Kunstler et al, 2022). Breathing discomfort was also identified as an adverse outcome that is potentially exacerbated by respirator use, though the evidence for this is also limited (Galanis et al, 2021; Kunstler et al, 2022).

Finally, multiple reviews note the impact of these adverse outcomes on adherence to PPE guidance overall. Issues with discomfort and fit potentially increase the risk of infection, especially in cases where itching occurs as an adverse outcome as there is the potential that any adjustment of the respirator or face covering may compromise the seal (Kunstler, et al 2022) and result in increased risk of infection.

#### 3.1.2 Barriers to PPE adherence

Evidence regarding the potential barriers to PPE use and adherence to guidelines is primarily explored in Houghton et al. (2020). The most commonly identified barrier across all literature included in Houghton et al. (2020) is ineffective or unclear communication in the workplace, especially where there are conflicting recommendations between local guidelines and national or international guidelines – this is reported in Houghton et al. (2020) and Gangaram et al (2022). Similar to this, workplace culture was identified as both a barrier and a facilitator for adherence in McCann-Pineo (2022).

Another barrier identified in the literature centers around the provision and quality of PPE that is available to HCWs. Many HCWs identified supply chain issues of N95 masks prior to the COVID-19 pandemic, which only worsened as the length of time PPE was required increased (Houghton et al, 2020). Additional barriers were identified at a more granular level by Houghton et al (2020) but were not included in this synthesis as they were not reported by any other sources used in this review.

#### 3.1.3 Facilitators to PPE adherence

Facilitators to PPE use were not explored in-depth in the literature. Houghton et al (2020) identified that a healthcare facility environment that had sufficient space for proper isolation of patients was regarded as a key facilitator for compliance with general IPAC guidelines, but this was specific to the hospital setting and is of limited relevance to this review. Individual beliefs and values were identified as a significant facilitator for adherence, being primarily centered on HCWs’ fears of contracting SARS or MERS resulting in increased vigilance. Similarly, concerns around transmission to family and co-workers, as well as a desire to provide high-quality care resulted in a strong sense of value for individual HCWs when adhering to IPAC guidelines.

### 3.2 Strengths and limitations of the available evidence

All studies included in this rapid review were formally assessed for risk of bias, the full process for which is outlined in section 6 of this review.

Following risk of bias assessment, the majority of the evidence available for this rapid review is of ‘Poor’ quality (5 studies). Caution should therefore be taken when drawing conclusions from the evidence. It is also important to note that many of the studies included in this review reflect data captured in a wide variety of healthcare settings and are not limited to our settings of interest – this raises implications for the generalisability of this review’s findings. We have mitigated this where possible by including only studies that have captured data from at least one of the settings of interest, but it is pertinent to note that the majority of the evidence is dominated by front-line hospital and nursing experiences.

### 3.3 Implications for policy and practice

This review has identified a paucity of evidence for the barriers, facilitators, and potential adverse effects of PPE usage in the settings of interest. While some of the findings from the literature included in this review may be generalizable to these settings, it is crucial that more high-quality research is undertaken in the areas of interest.

### 3.4 Strengths and limitations of this Rapid Review

All studies included in this rapid review were identified by systematically reviewing a range of carefully selected publication databases, as outlined in Appendix 1. The research question and study protocol were developed in collaboration with external shareholders and experts in the field. The abstract, full text screening and data extraction were performed by a single reviewer and checked for consistency by a second researcher. The methods used in this review have been robust and pragmatic, but due to the nature of rapid reviews there remains the possibility that additional eligible texts were not identified.

This rapid review is strictly limited to a select few studies that align with the research question and protocol. We are reliant on interpreting the results of studies that have several limitations and differing levels of quality, and this impacts the strength of our conclusions. We have summarised the primary limitations of studies included in this review, and each included study has been assessed for risk of bias.

## Data Availability

All data produced in the present study are available upon reasonable request to the authors

## Abbreviations

Acronym: Full Description
IPAC: Infection Prevention and Control
PPE: Personal Protective Equipment
EMS: Emergency Medical Services
GP: General Practice / Practitioner(s)
AGPs: Aerosol-Generating Procedures
HMCAS: Hamad Medical Corporation Ambulance Service
RCT: Randomised Controlled Trial
HCW: Healthcare Worker(s)
RoB: Risk of Bias
ROBIS: Risk of Bias in Systematic Reviews
MERS-CoV: Middle Eastern Respiratory Syndrome
SARS: Severe acute respiratory syndrome
H1N1: H1N1 strain of the flu (influenza A) virus

## 5. RAPID REVIEW METHODS

### 5.1 Eligibility criteria

### 5.2 Literature search

The search strategy was developed by Health Technology Wales and shared with members of the WCEC. As the number of hits returned for this topic are large, various ways to conduct the search were investigated. It was decided to restrict the search to the population (COVID-19 and related respiratory viruses) and the intervention (personal protective equipment (PPE)).

No attempt was made to search by setting, due to the inconsistent way that setting is reported in the literature. The decision was taken, therefore, to separate the results into evidence ‘layers’ using study design filters. Each layer will be mutually exclusive:

- Secondary evidence
- Primary evidence
- Qualitative evidence
- Other (i.e. anything that did not fit into the first three layers)

Two versions of the PPE stem were developed in Medline; the second of which was a focussed version of the first. This aimed to reduce the number of hits and only retrieve those which pertained to PPE, as opposed to a passing reference in the abstract. Following a comparison of the two PPE stems when combined with the virus stem (the COVID-19 inbuilt filter within Medline and a stem for the other related respiratory viruses) it was decided to use of the focussed PPE stem for the search.

As the work includes respiratory viruses in addition to COVID-19, all COVID-19 specific databases were excluded from the search as all sources searched should include content pertaining to all viruses of interest, thereby ensuring no unfair weighting of the results. Also, the search methods employed to focus the PPE search make use of complex searching functionality which is not available in the COVID-19 specific resources (and some other resources). Searching these would then undo any benefits gained through the focussing of the PPE stem. Registers of clinical trials and systematic reviews were also searched.

The search strategy is available in Appendix 2.

### 5.3 Study selection process

Study screening and selection against the eligibility criteria was carried out by Antonia Needham, with selection decisions checked by Lauren Elston and any disagreements resolved by consensus amongst the researchers. After sifting of the first three evidence layers, it was decided by consensus that the fourth layer of evidence (detailed in 5.2) would not be used due to a sufficient amount of available evidence in the first three layers.

### 5.4 Data extraction

Data was extracted as documented in Tables 1 and 2 by Antonia Needham. Extraction was checked by Lauren Elston.

**Table 1:**
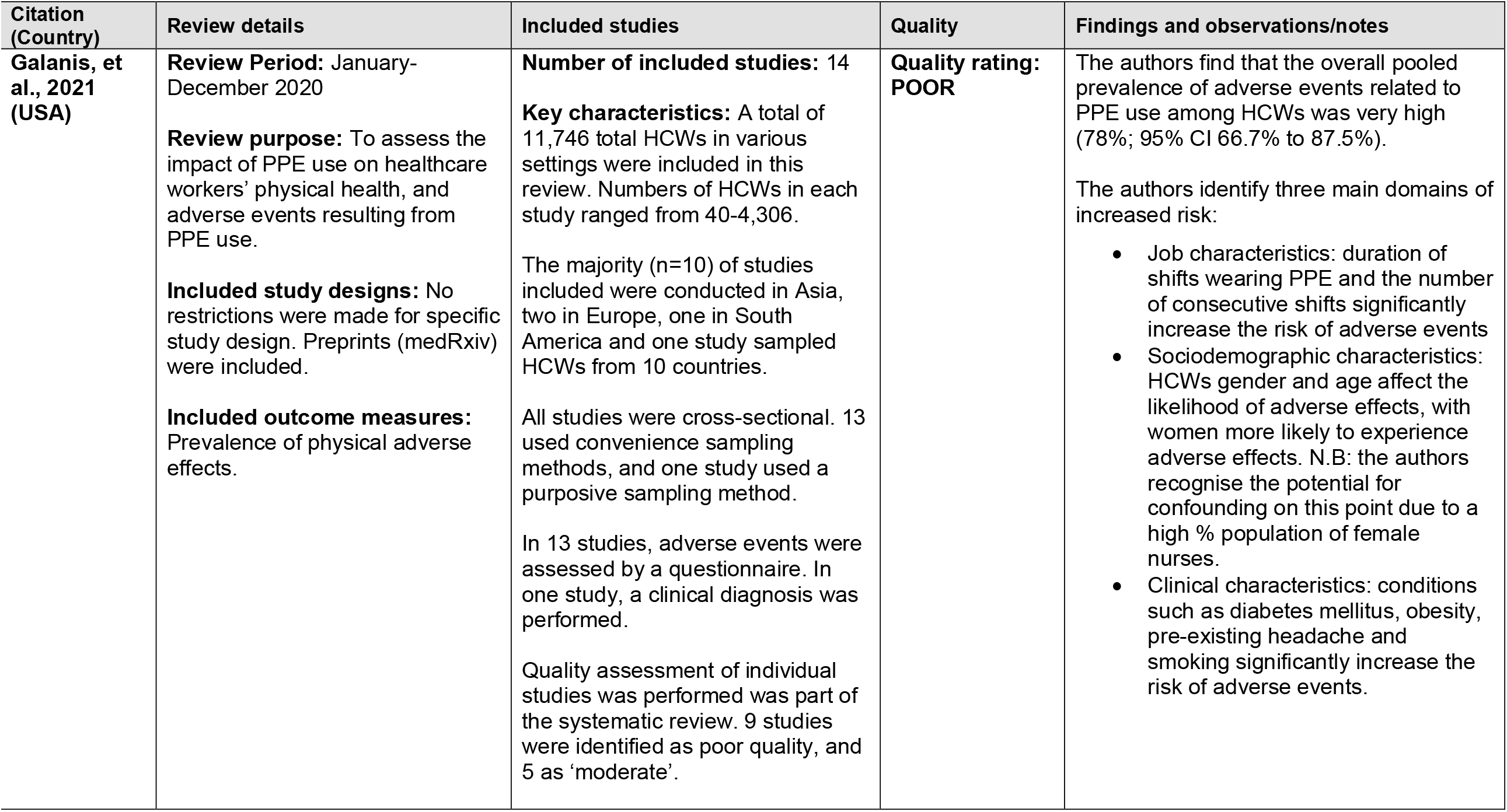
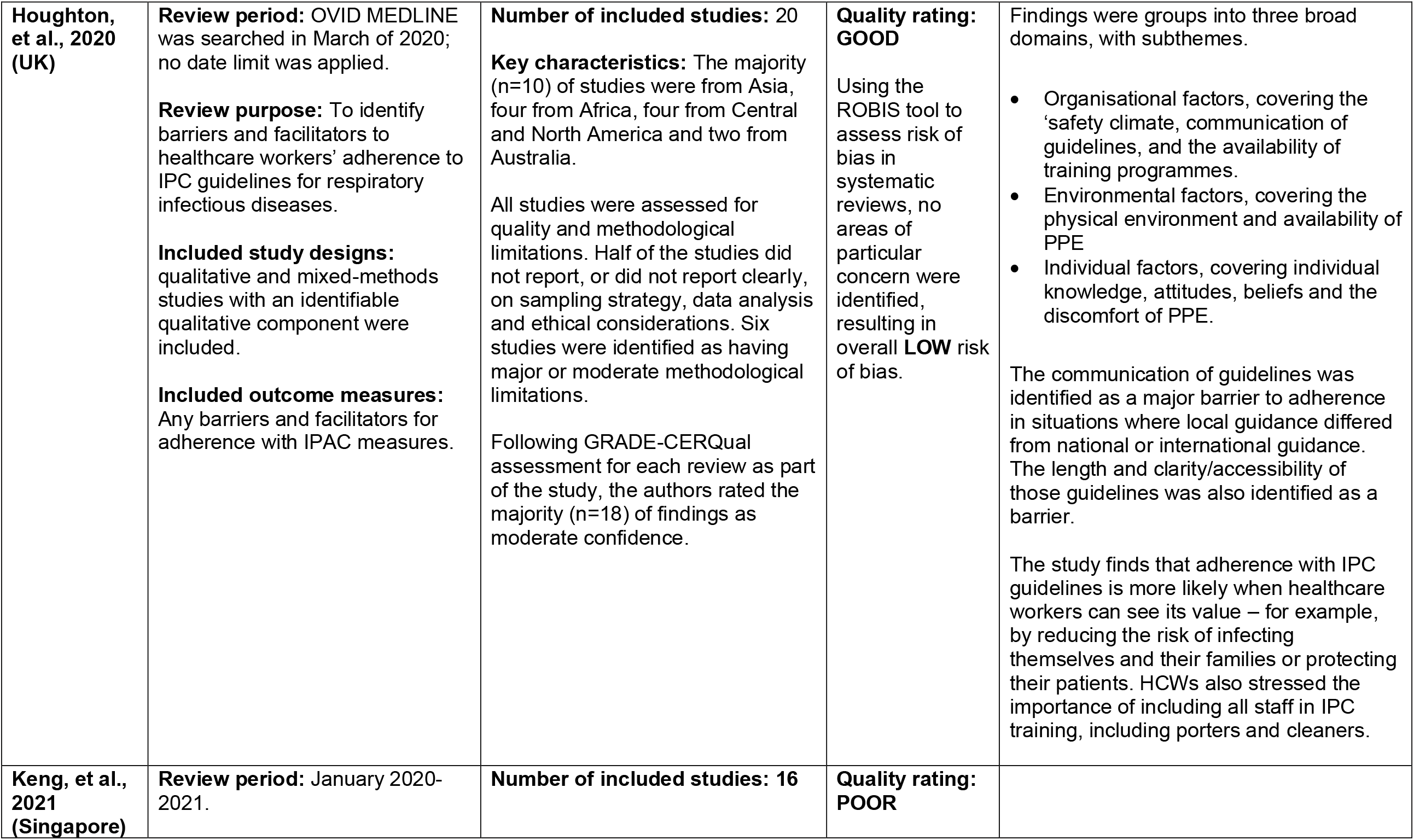
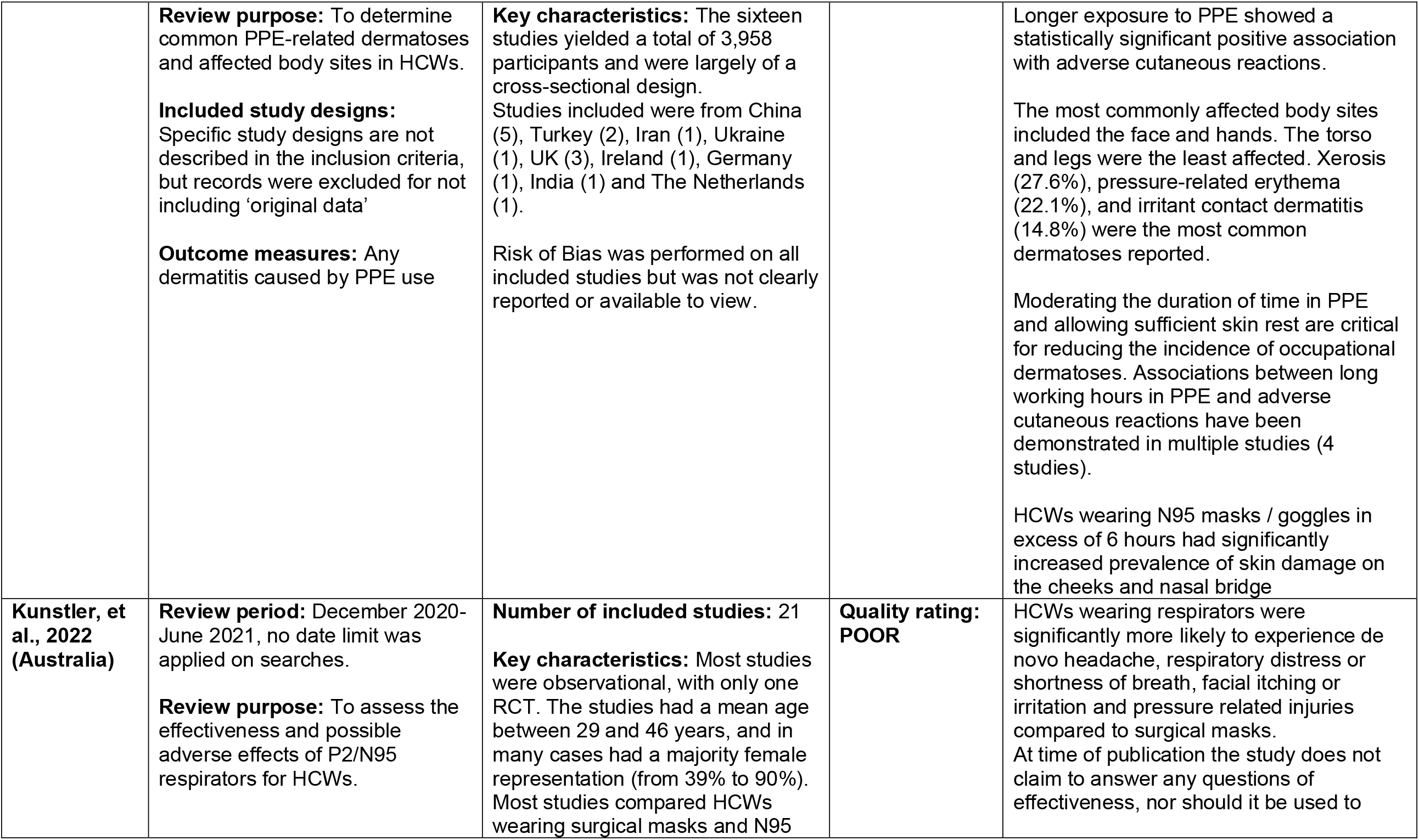
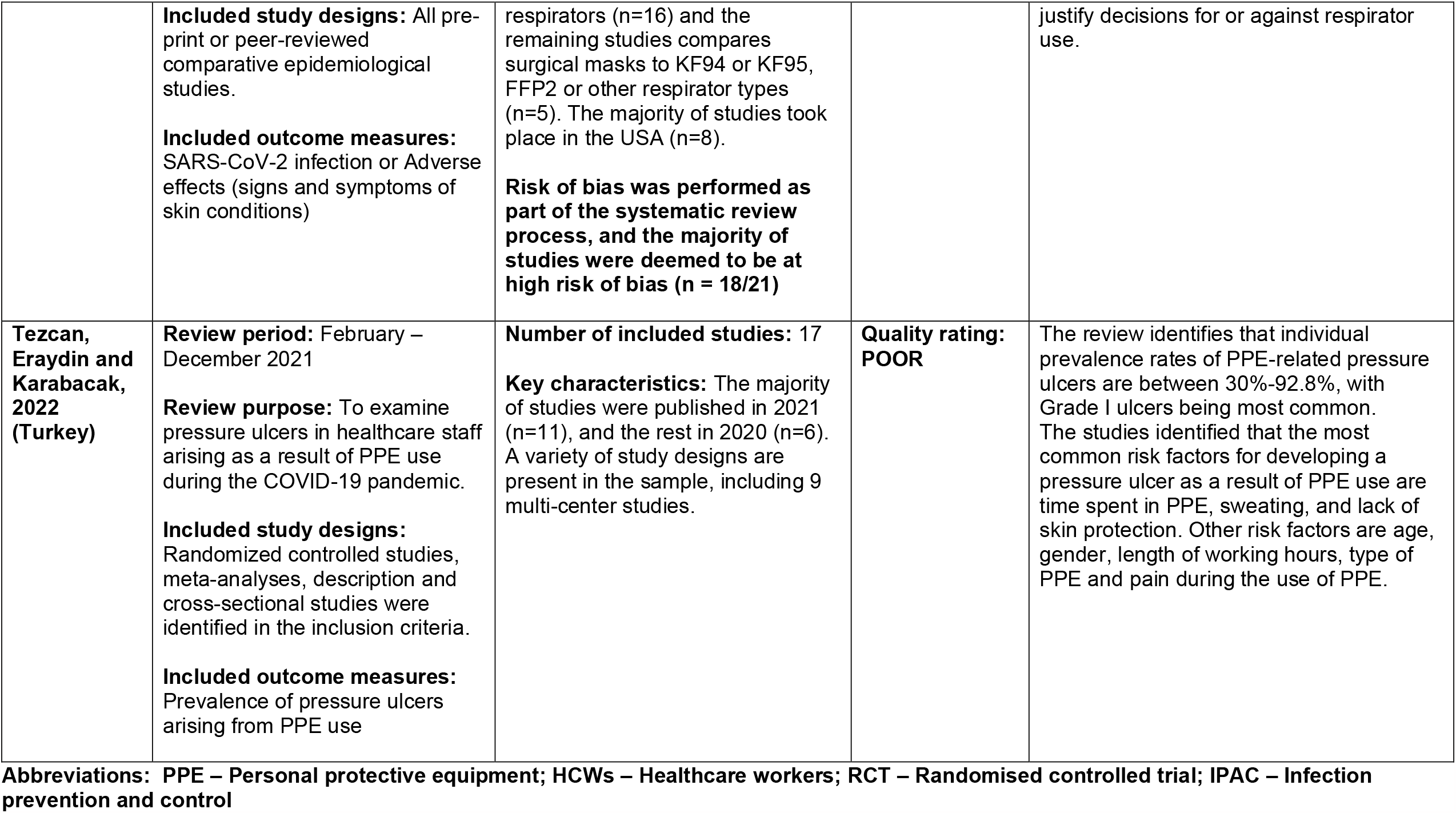
Summary of Systematic Reviews

**Table 2:** Summary of Primary Studies

| **Citation (Country)** | **Study Details** | **Participants & setting** | **Key findings** | **Observations/notes** |
| --- | --- | --- | --- | --- |
| **Gangaram, Pillay and Alinier., 2022 (Qatar)** | **Study Design:** Prospective quantitative cohort study.<br>**Type of intervention [exposure]:** Use of PPE<br>**Data collection methods:** Online survey<br>**Quality rating: FAIR** | **Sample size:** 282<br>**Participants:** Paramedics employed by the Hamad Medical Corporation Ambulance Service (HMCAS)<br>**Setting:** Ambulance<br>**Dates of data collection:** July 2020 | **Primary Findings:** 98.2% of surveyed paramedics are knowledgeable about methods of transmission for COVID-19. Regular completion of PPE training was successful in embedding infection control attitudes.<br>**Additional Findings:** 63% of surveyed paramedics agree the donning and doffing of PPE may be more effective when supervised. | Participant characteristics were not reported.<br>The survey was stopped as soon as the sample size (determined by Slovin's formula) was reached. |
| **McCann-Pinneo, Levinsky and Berkowitz. 2022. (USA)** | **Study Design:** Medical record review.<br>**Type of intervention [exposure]:** Use of PPE<br>**Data collection methods:** Retrospective chart review among all EMS encounters for a New York health system.<br>**Quality rating: POOR** | **Sample size:** 28,693 EMS encounters<br>**Participants:** EMS responders affiliated with a large New York health system<br>**Setting:** Emergency medical services / Ambulance<br>**Dates of data collection:** March-June 2020 | **Primary Findings:** Full PPE utilization was most strongly influenced by dispatch codes that were indicative of breathing problems and respiratory arrest, in addition to a patient's positive screening for COVID-19 on dispatch. Partial PPE use was documented in 92.8% of encounters, full use in 17.8%. | Retrospective study design and the use of chart reviews may mean results are reported inconsistently. Some EMS calls were excluded due to lack of data. |

**Abbreviations:** HMCAS – Hamad Medical Corporation Ambulance Service; EMS – Emergency medical services

**Table 3:** Eligibility criteria

|  | **Inclusion criteria** | **Exclusion criteria** |
| --- | --- | --- |
| **Participants** | All staff |  |
| **Settings** | Emergency ambulance settings<br>General practice, including district nurse care | Any other primary care or community care setting (e.g. Dental, Optometry, Domiciliary and Care Home, Pharmacy)<br>Any secondary or tertiary settings |
| **Intervention / exposure** | Personal protective equipment (PPE) for COVID-19 and related respiratory viruses | Other infection prevention and control (IPAC) measures<br>Other non-pharmaceutical interventions (NPIs) |
| **Comparison** | No PPE measures in place<br>Other PPE measures |  |
| **Outcomes** | User preference (factors affecting adherence, possible harms, barriers and facilitators for use)<br>Quality of life impacts |  |
| **Study design** | We will search and include evidence in the following order<br>Systematic reviews, rapid reviews<br>Primary evidence and qualitative evidence |  |
| **Countries** | Any |  |
| **Language of publication** | English |  |
| **Publication date** | 2020-2022 |  |
| **Publication type** | Published and preprint |  |
| **Reporting of data** | We will consider other study designs where their findings may be useful for providing context to the review (e.g. narrative reviews) |  |

### 5.7 Quality appraisal

Formal quality assessments were completed depending on applicability and study design. All studies were formally assessed for their risk of bias. The five secondary sources were assessed using the Risk of Bias in Systematic Reviews (ROBIS) tool. Both primary studies were assessed using the NIH quality assessment tool for observational and cross-sectional studies. One study (Gangaram et al, 2022) was also assessed using the critical appraisal skills programme (CASP) checklist for qualitative research, due to difficulties finding an appropriate risk of bias assessment tool for the study design. For this study, only the NIH results are reported.

### 5.8 Synthesis

Quantitative analysis of relevant outcomes was not possible for this rapid review. As a result, evidence was synthesised narratively in relevant sections and individual outcome data are listed in relevant tables within this review.

## 6. EVIDENCE

### 6.1 Study selection flow chart

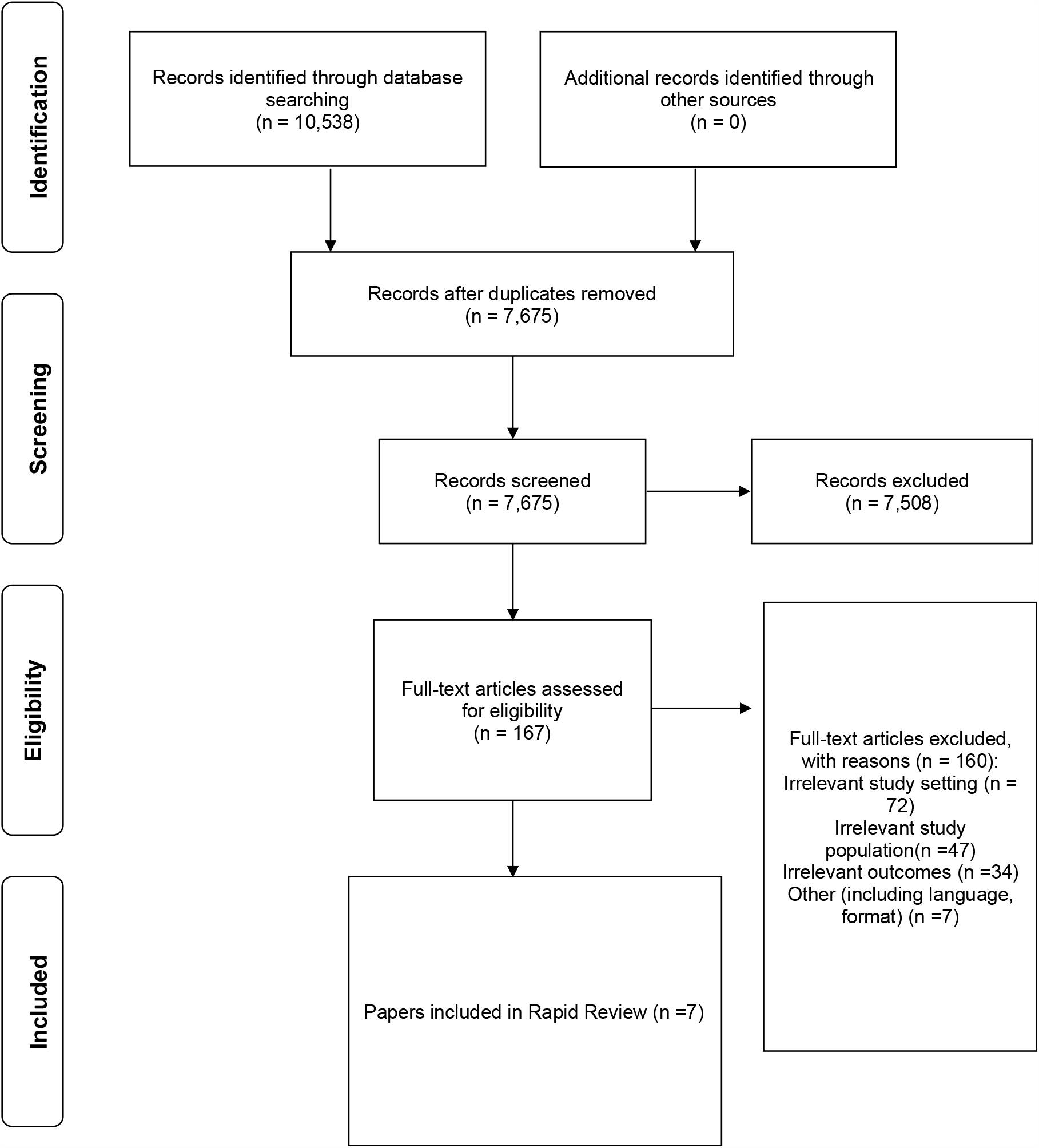

### 6.3 Quality appraisal tables

The risk of bias (RoB) tools used for this review are outlined in section 5.7. Risk of bias tools assess studies across different domains to ascertain whether there is a potential that the evidence is biased.

| Reference | RoB tool used | Overall RoB | Comments |
| --- | --- | --- | --- |
| **Galanis et al (2021)** | ROBIS (Whiting et al. 2016) | **HIGH** | Reviewers did not address uncertainty around search strategy, characteristics that could influence outcomes. |
| **Houghton et al (2020)** | ROBIS (Whiting et al. 2016) | **LOW** | No major concerns identified, authors reflected on their role in analysis and limitations were discussed |
| **Keng et al (2021)** | ROBIS (Whiting et al. 2016) | **HIGH** | Serious omissions or limitations in the approach taken by the review. No discussion of limitations or reflection on validity of evidence. No statistical analysis |
| **Kunstler et al (2022)** | ROBIS (Whiting et al. 2016) | **HIGH** | Concerns relating to heterogeneity were not addressed by the authors, relevance was not discussed within the review. |
| **Tezcan et al (2022)** | ROBIS (Whiting et al. 2016) | **HIGH** | Limitations were not discussed within the review, concerns relating to formal risk of bias and search strategy. |
| **Gangaram et al (2022)** | NIH quality assessment tool for observational and cross-sectional studies | **UNCLEAR** | Primary concerns relate to participant identification and whether using power number as a stopping criteria was appropriate. NIH tool was also partially suitable for this study – CASP was also used to assess and is available upon request. |
| **McCann-Pineo et al (2022)** | NIH quality assessment tool for observational and cross-sectional studies | **HIGH** | Primary concerns centre around the retrospective study design and reliance of chart reviews, which are known for inconsistent reporting. Some EMS calls were excluded due to lack of data, and some characteristics not included in the regression without adequate explanation. |

### 6.2 Information available on request

The study protocol, full search strategy and list of excluded studies are available upon request.

## 7.2 Acknowledgements

The authors would like to thank Rowena Christmas, Nigel Rees, Louise Colson, Nathan Davies, Sue Tranka, Helen Arthur, Rob Orford, Emma Coles and Chris Jones for their time, contributions and expertise during stakeholder meetings, in guiding the focus of the review and interpretation of findings, and for reviewing the report.

## 8. ABOUT THE WALES COVID-19 EVIDENCE CENTRE (WCEC)

The WCEC integrates with worldwide efforts to synthesise and mobilise knowledge from research.

We operate with a core team as part of <u>Health and Care Research Wales</u>, are hosted in the <u>Wales Centre for Primary and Emergency Care Research (PRIME)</u>, and are led by <u>Professor Adrian Edwards of Cardiff University</u>.

The core team of the centre works closely with collaborating partners in <u>Health Technology Wales, Wales Centre for Evidence-Based Care, Specialist Unit for Review Evidence centre, SAIL Databank, Bangor Institute for Health & Medical Research/ Health and Care Economics Cymru</u>, and the <u>Public Health Wales Observatory</u>.

Together we aim to provide around 50 reviews per year, answering the priority questions for policy and practice in Wales as we meet the demands of the pandemic and its impacts.

### Director

Professor Adrian Edwards

### Website

https://healthandcareresearchwales.org/about-research-community/wales-covid-19-evidence-centre

## APPENDIX

**APPENDIX 1:** Resources searched during Rapid Review Searching.

| Database Searched | Date Searched | Evidence layer | No of Hits Retrieved |
| --- | --- | --- | --- |
| Ovid Medline (ALL) | 22/11/22 | Secondary | 652 |
|  |  | Primary | 1414 |
|  |  | Qualitative | 1241 |
|  |  | Other | 3828 |
| Ovid Embase | 22/11/22 | Secondary | 940 |
|  |  | Primary | 1235 |
|  |  | Qualitative | 1457 |
|  |  | Other | 3605 |
| Cochrane Library | 23/11/22 | Secondary | 14 |
|  |  | Primary | 465 |
| Epistemonikos | 22/11/22 | Secondary | 81 |
|  |  | Primary | 1802 |
| Trip Pro | 22/11/22 | Secondary | 130 |
|  |  | Primary | 132 |
| INAHTA HTA | 24/11/22 | Secondary | 7 |
| WHO ICTRP | 02/11/22 | ongoing studies | 126 |
| clinicaltrials.gov | 01/12/22 | ongoing studies | 455 |
| PROSPERO | 02/12/22 | ongoing studies | 387 |

**APPENDIX 2:**
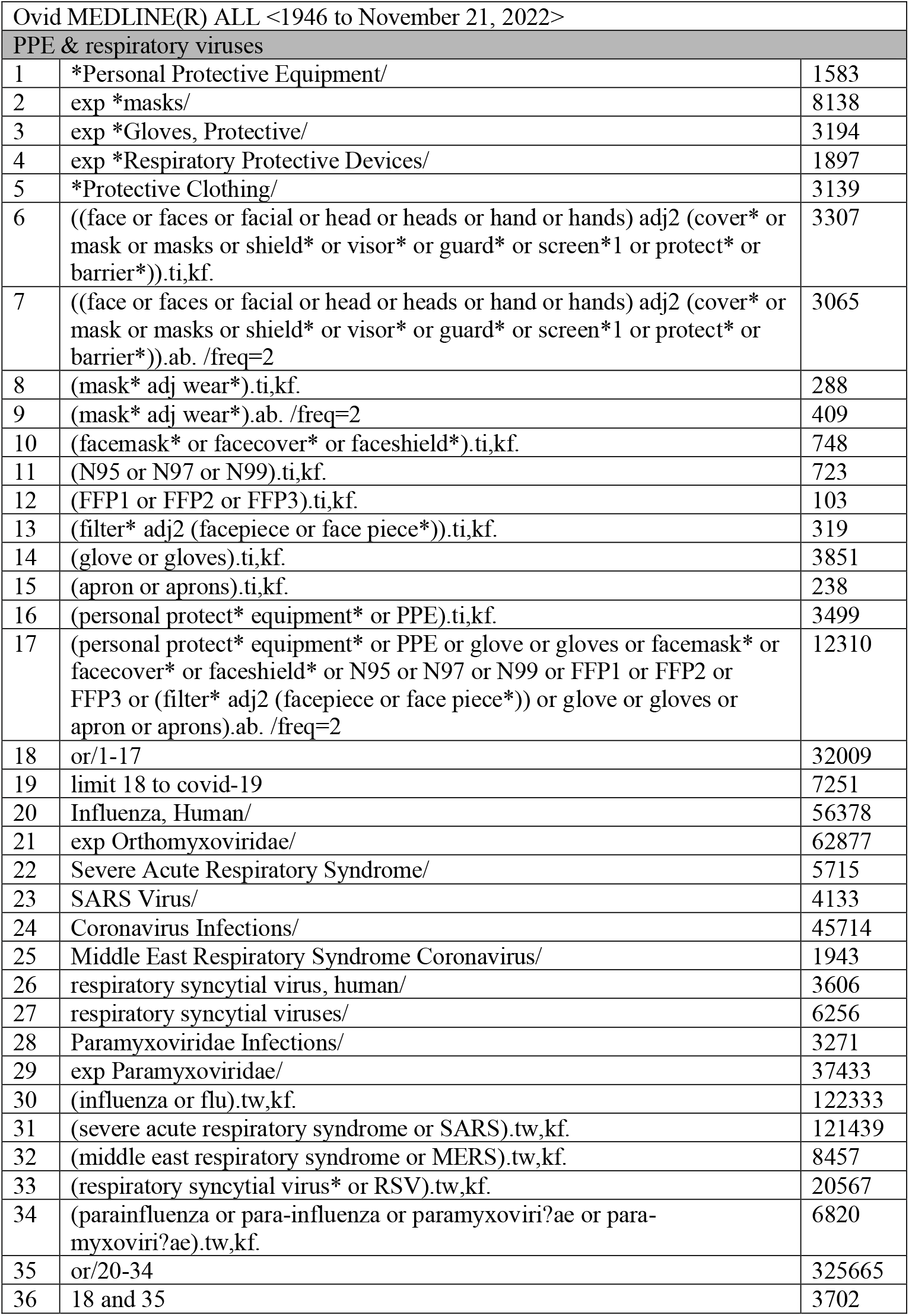
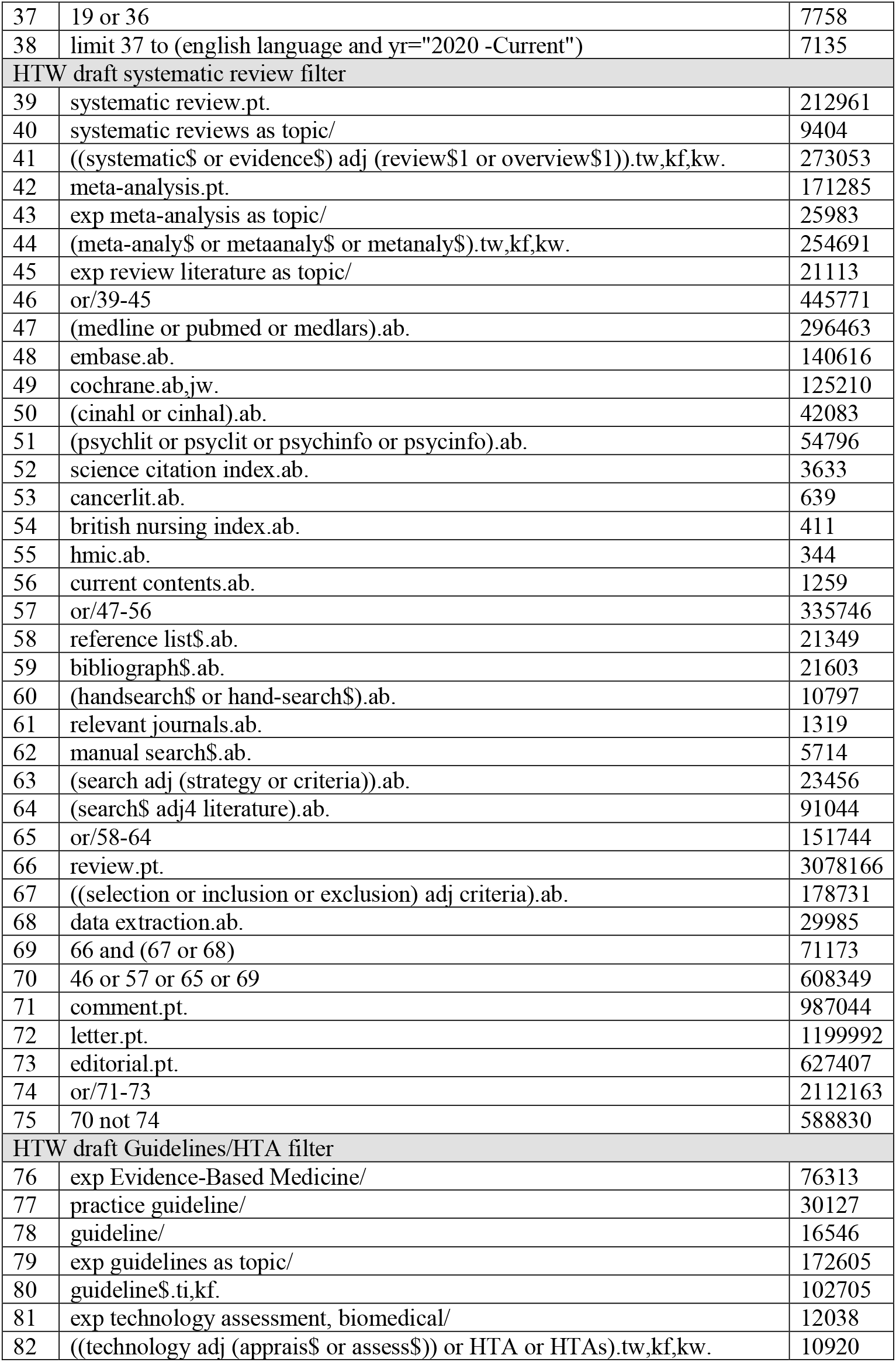
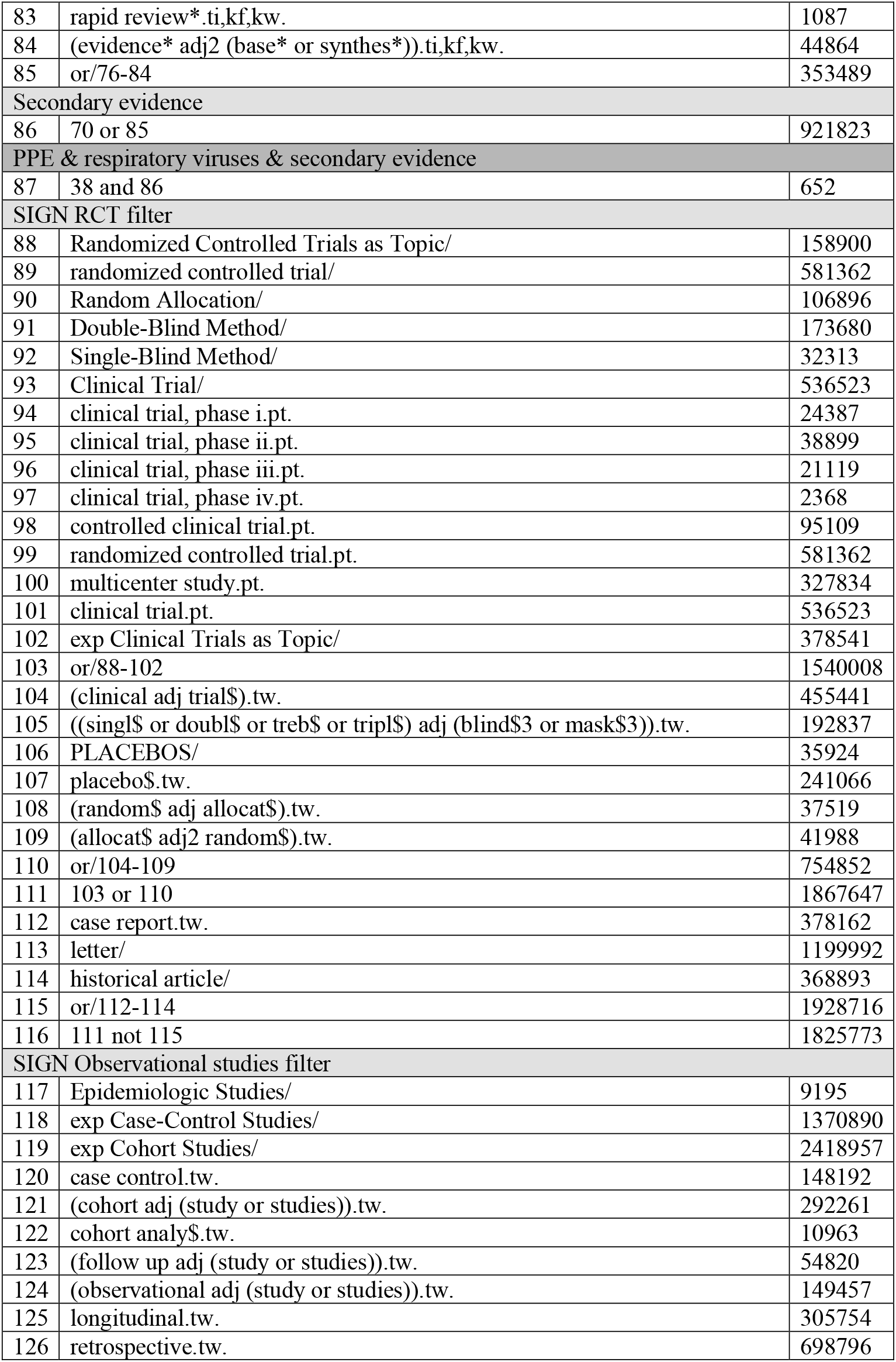
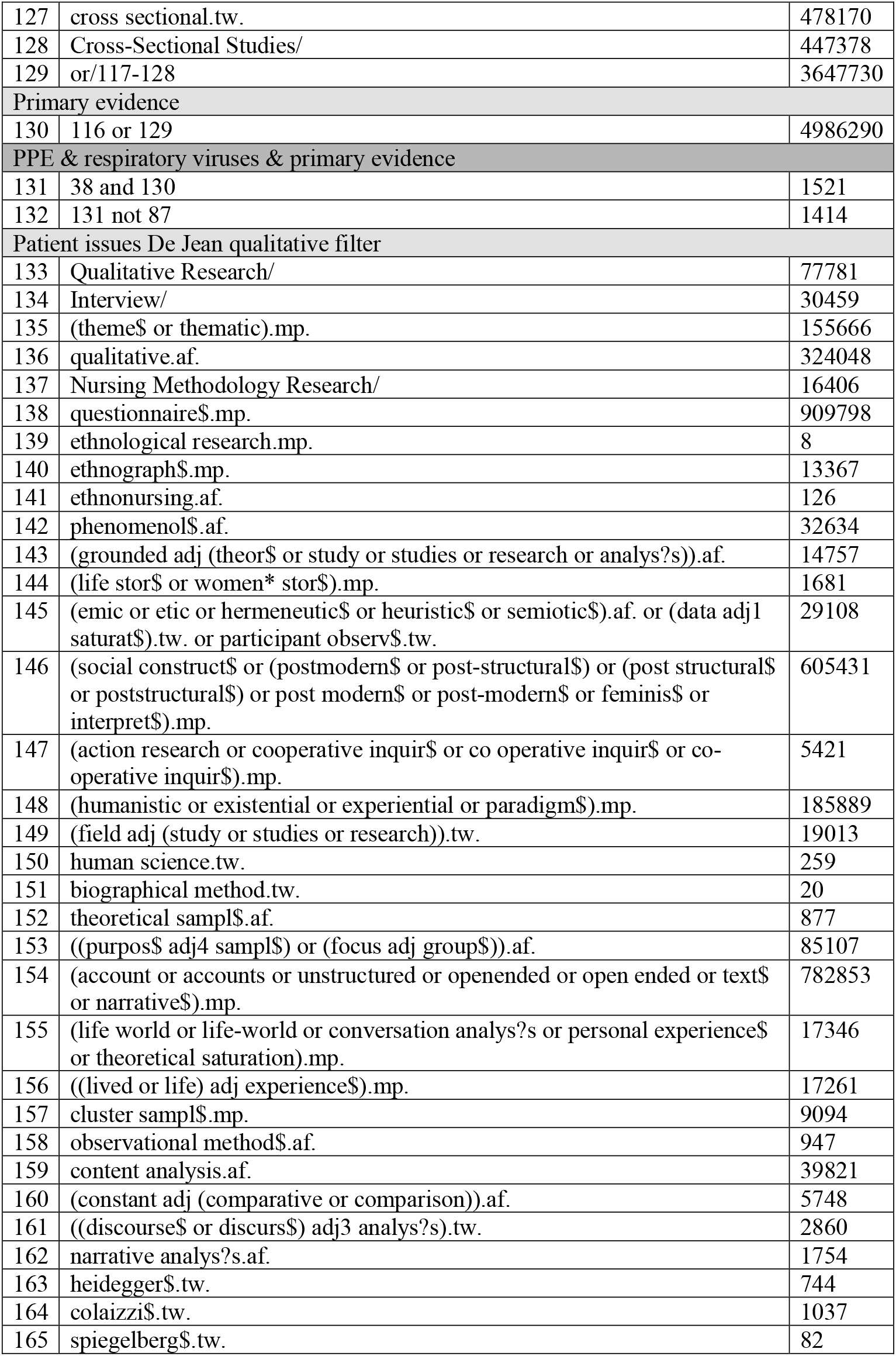
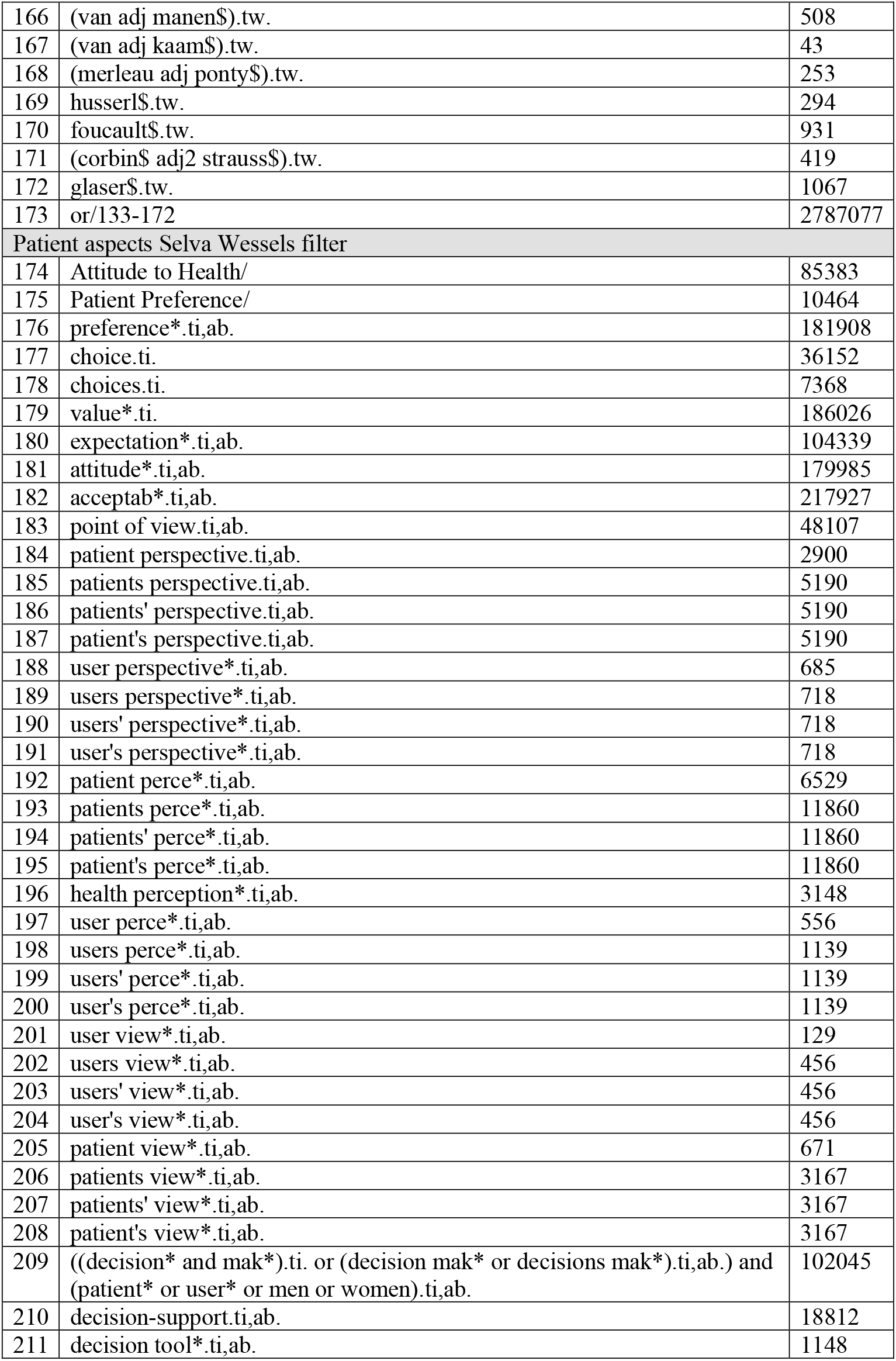
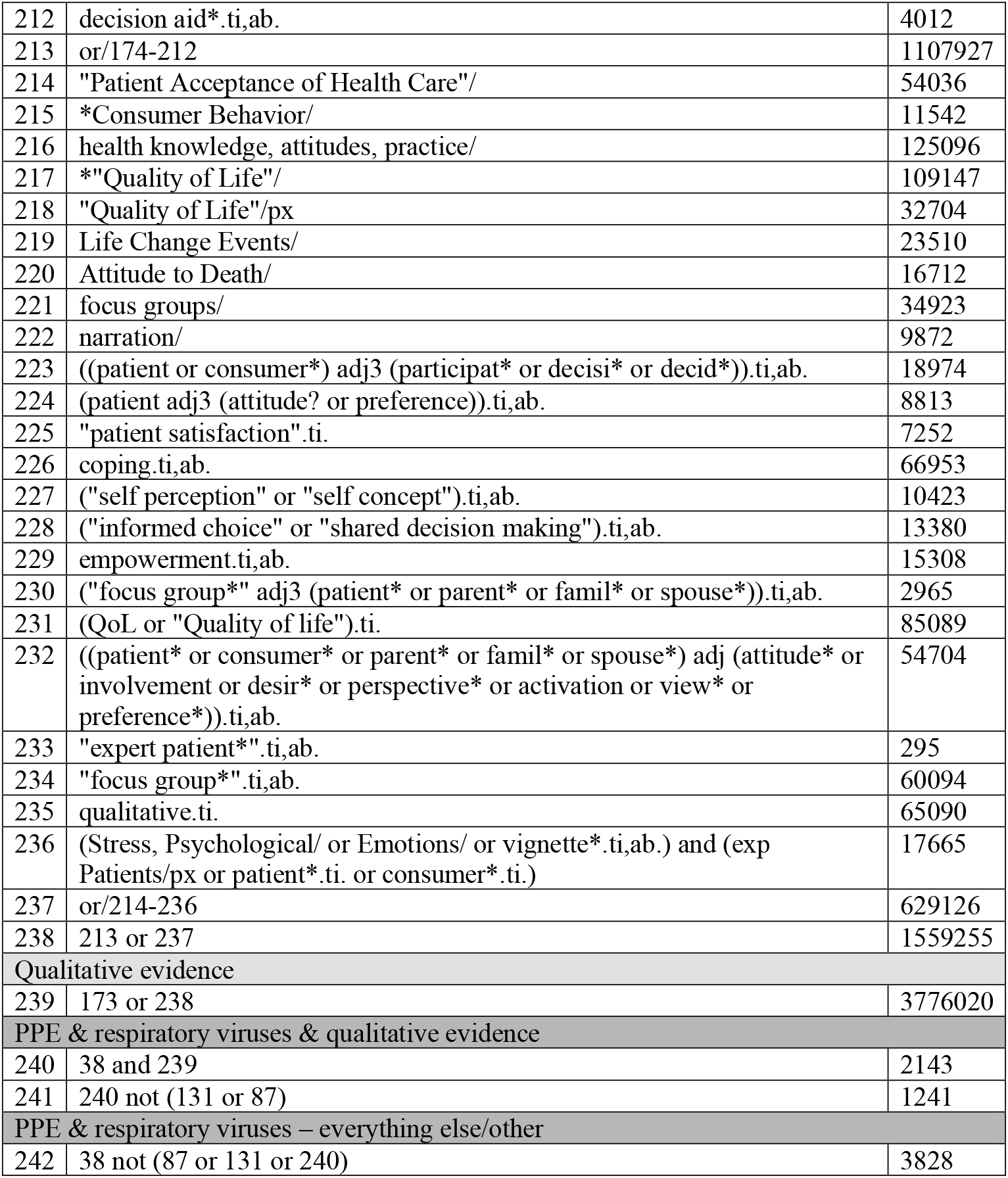
MEDLINE strategy.

